# Care Fragmentation and Outpatient Imaging Utilization and Expenditures in U.S. Adults: A MEPS 2020 Study

**DOI:** 10.1101/2025.08.23.25334292

**Authors:** Andrew Bouras, Dhruv Patel

## Abstract

**Background:** Care fragmentation has been linked to overuse and higher costs. Evidence on how fragmentation relates to outpatient imaging in the general U.S. population remains limited.

**Objective:** To quantify the association between care fragmentation and outpatient imaging utilization and radiology expenditures among U.S. adults, and to test whether associations differ by multimorbidity and insurance.

**Design:** Cross-sectional, survey-weighted analysis using the Medical Expenditure Panel Survey (MEPS) 2020.

**Participants:** Adults aged *≥*18 years in MEPS 2020 with non-missing survey design variables.

**Exposures:** Care fragmentation category (None/Low/Medium/High), defined using person-level utilization breadth and intensity from the Full-Year Consolidated file.

**Main outcomes and measures:** Primary outcomes included any advanced imaging (receipt of at least one MRI, CT, or ultrasound during office-based visits) and total radiology expenditures (sum of payments for imaging-flagged office-based visits). Secondary outcomes included any imaging, imaging visit count, and radiology spending as a proportion of total expenditures.

**Results:** In survey-weighted models (reference = Low; “None” excluded from regressions), Medium fragmentation was associated with higher odds of any advanced imaging (OR 1.42, 95% CI 1.23–1.64), whereas High did not further increase the probability (OR 0.92, 0.71–1.19). Fragmentation substantially increased intensity: imaging visit counts (RR 2.51 Medium; 3.96 High vs Low) and radiology spending among spenders (2.36× Medium; 3.96× High vs Low). Older age (50–64, 65+), female sex, and Medicare coverage independently raised both imaging probability and spend.

**Conclusions:** Care fragmentation is associated with both increased probability and intensity of advanced imaging utilization, with particularly strong effects on radiology expenditures among adults with positive spending. Findings support targeted interventions to improve care coordination and reduce potentially unnecessary imaging in fragmented care environments.

## 1 Introduction

Healthcare fragmentation, defined as receiving care across multiple providers and settings without adequate coordination or continuity, has emerged as a significant concern in contemporary healthcare delivery. Prior research has demonstrated that fragmented care is associated with increased healthcare utilization, including more diagnostic testing, procedures, emergency department visits, and hospitalizations[2, 1]. At the population level, care fragmentation has been linked to potentially inappropriate medication use, preventable complications, and increased mortality, particularly among patients with chronic conditions[3, 14].

Advanced imaging utilization, including magnetic resonance imaging (MRI), computed tomography (CT), and ultrasound, represents a substantial component of healthcare spending and has been subject to considerable policy attention due to concerns about overuse and cost containment[5, 6]. National trends indicate that advanced imaging utilization grew rapidly during the early 2000s before moderating in response to policy interventions and increased cost-sharing, with notable variations across demographic groups and insurance types[7, 8, 9]. The Medicare population, in particular, has shown continued growth in imaging utilization despite overall moderation in growth rates.

The theoretical relationship between care fragmentation and imaging utilization is supported by several mechanisms. Fragmented care may lead to information gaps between providers, resulting in duplicative testing when imaging results are not readily available or adequately communicated[10, 11]. Additionally, providers in fragmented care environments may practice more defensive medicine, ordering additional imaging studies to compensate for incomplete clinical information or uncertainty about previous testing[12]. Patients receiving care from multiple specialists may also experience higher imaging utilization as each provider orders tests within their specialty domain without full knowledge of imaging already performed by other providers.

Despite the theoretical rationale and growing policy concern about both care fragmentation and imaging overuse, empirical evidence examining the relationship between fragmentation and outpatient imaging utilization in the general U.S. population remains limited. Most existing studies have focused on Medicare beneficiaries or specific patient populations, leaving gaps in our understanding of how fragmentation affects imaging use across the broader adult population with diverse insurance coverage and health status[1, 15]. Furthermore, previous studies have often examined utilization patterns without corresponding analysis of expenditure effects, limiting understanding of the economic implications of fragmentation-related imaging use.

Understanding the relationship between care fragmentation and imaging utilization has important implications for health policy and clinical practice. If fragmentation contributes to unnecessary imaging, targeted interventions to improve care coordination could simultaneously improve quality and reduce costs. Conversely, if fragmentation-related imaging reflects appropriate care for complex patients requiring multiple specialists, policies to reduce fragmentation might inadvertently limit access to needed services.

Therefore, we conducted this study to quantify the association between care fragmentation and outpatient imaging utilization and radiology expenditures among U.S. adults using nationally representative survey data. We hypothesized that higher levels of care fragmentation would be associated with increased probability of receiving advanced imaging and higher radiology expenditures. We further examined whether these associations varied by multimorbidity status and insurance type, given that these factors may modify both fragmentation patterns and imaging needs.

## 2 Methods

### 2.1 Data source

We analyzed data from the Medical Expenditure Panel Survey (MEPS) 2020, a nationally representative survey of the U.S. civilian noninstitutionalized population conducted by the Agency for Healthcare Research and Quality. Our analysis utilized four public-use files: the Full-Year Consolidated file (FYC; HC-224) containing demographic, socioeconomic, and annual utilization summaries; the Office-Based Medical Provider Visits file (HC-220G) with detailed visit-level information including service indicators; the Condition–Event Link file (HC-220IF1) connecting medical conditions to healthcare events; and the Medical Conditions file (HC-222) containing diagnosed conditions with clinical classification codes. All analyses incorporated the complex survey design elements specified in HC-224 documentation, including person-level weights, variance strata, and primary sampling units.

### 2.2 Population

Our study population included adults aged 18 years and older from the MEPS 2020 sample with complete survey design variables (person-level weight PERWT20F, variance stratum VARSTR, and primary sampling unit VARPSU). We excluded respondents with missing survey design variables to ensure appropriate variance estimation. The final analytic sample comprised 22,339 adults representing approximately 251.4 million U.S. adults when weighted. Sample size calculations indicated adequate power (*>*80%) to detect odds ratios of 1.3 or greater for primary outcomes, assuming *α* = 0.05 and accounting for the complex survey design effect.

### 2.3 Variables

Our primary exposure was care fragmentation, operationalized as a four-category variable (None/Low/Medium/High) based on breadth and intensity of healthcare utilization patterns derived from the Full-Year Consolidated file. We defined fragmentation categories using a composite measure incorporating office-based visits, emergency room visits, and inpatient stays. The *None* category included individuals with no recorded office-based, emergency room, or inpatient encounters. *Low* fragmentation was defined as 1–4 office-based visits with no emergency room or inpatient stays. *Medium* fragmentation included 5–9 office-based visits or any combination with 1 emergency room visit but no inpatient stays. *High* fragmentation encompassed *≥*10 office-based visits, multiple emergency room visits, or any inpatient stays. These cutpoints were established a priori based on clinical interpretability, prior literature on healthcare utilization patterns, and distributional considerations to ensure adequate sample sizes across categories while maintaining meaningful distinctions in care complexity.

The primary outcomes included any advanced imaging (defined as MRI, CT, or ultrasound) received during office-based visits and total radiology expenditures (calculated as the sum of payments for imaging-flagged office-based visits). Secondary outcomes encompassed any imaging (including X-ray and mammography), imaging visit count, and radiology spending as a proportion of total healthcare expenditures. We examined two key effect modifiers: multimorbidity (defined as having two or more chronic conditions based on HC-222 condition records) and insurance type (categorized as Medicare plus Medicaid dual coverage, Medicare only, Medicaid only, private insurance, or uninsured/other coverage). Covariates included age group, sex, race/ethnicity, poverty status, census region, and total number of office visits to control for overall healthcare utilization.

### 2.4 Imaging ascertainment

Imaging utilization was ascertained from the HC-220G Office-Based Medical Provider Visits file using visit-level service indicators for X-ray (XRAYS_M18), MRI (MRI_M18), ultrasound/sonogram (SONOGRAM_M18), and mammogram (MAMMOG_M18). Advanced imaging was defined as any MRI, CT, or ultrasound procedure, excluding routine X-rays and mammograms to focus on higher-cost, potentially discretionary imaging. We aggregated visit-level indicators to create person-year binary outcomes (any advanced imaging, any imaging overall) and count outcomes (number of imaging visits). Radiology expenditures were calculated as the sum of total payments (OBXP20X) across all office-based visits flagged as having imaging services, representing both out-of-pocket and third-party payments for imaging-related care.

### 2.5 Statistical analysis

We implemented survey-weighted analyses using the complex survey design specified in MEPS documentation, constructing the survey design object with primary sampling units (VARPSU), strata (VARSTR), and person-level weights (PERWT20F) with nested sampling structure. For the primary outcome of any advanced imaging, we employed survey-weighted logistic regression models with fragmentation category as the main exposure, adjusting for age group, sex, race/ethnicity, poverty status, insurance type, and total office visits. We tested for interactions between fragmentation and both multimorbidity status and insurance type using likelihood ratio tests. For radiology expenditures, we used a two-part modeling approach to address the high proportion of zero spending: the first part employed logistic regression to model the probability of any radiology spending, while the second part used generalized linear models with gamma family and log link to model spending amounts among those with positive expenditures. We conducted sensitivity analyses including alternative fragmentation cutpoints, restricting advanced imaging to MRI and CT only (excluding ultrasound), and using provider specialty as a proxy for radiology services when imaging flags were missing. All analyses incorporated Taylor series linearization for variance estimation and used appropriate survey procedures to account for the complex sampling design.

### 2.6 Reporting

We present results following STROBE guidelines for cross-sectional studies. Figure 1 displays predicted probabilities of receiving advanced imaging across fragmentation levels, stratified by multimorbidity status with 95% confidence intervals. Figure 2 shows survey-weighted mean radiology expenditures across fragmentation categories, stratified by insurance type with corresponding confidence intervals.

**Figure 1.**
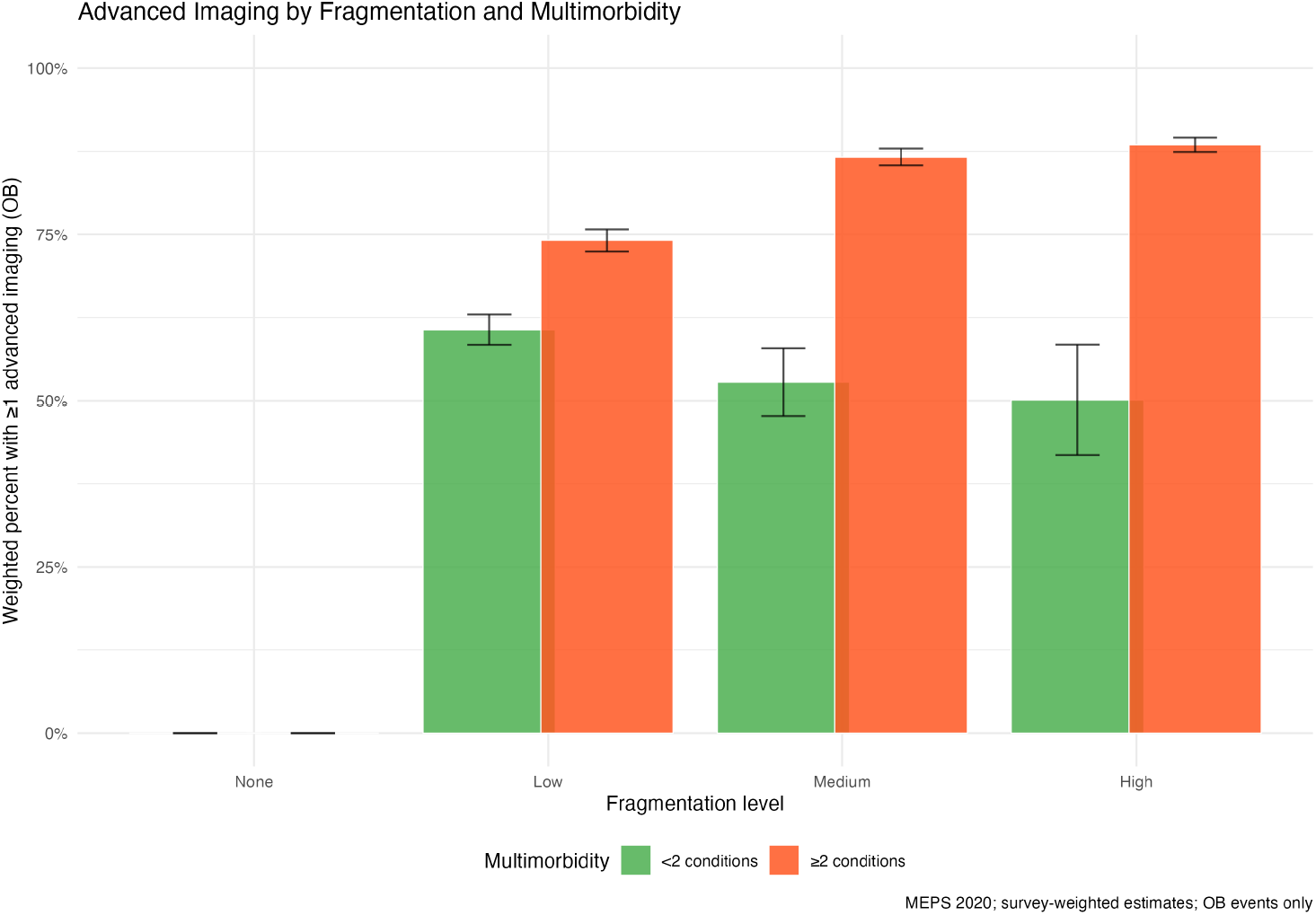
Advanced imaging by fragmentation and multimorbidity.

**Figure 2.**
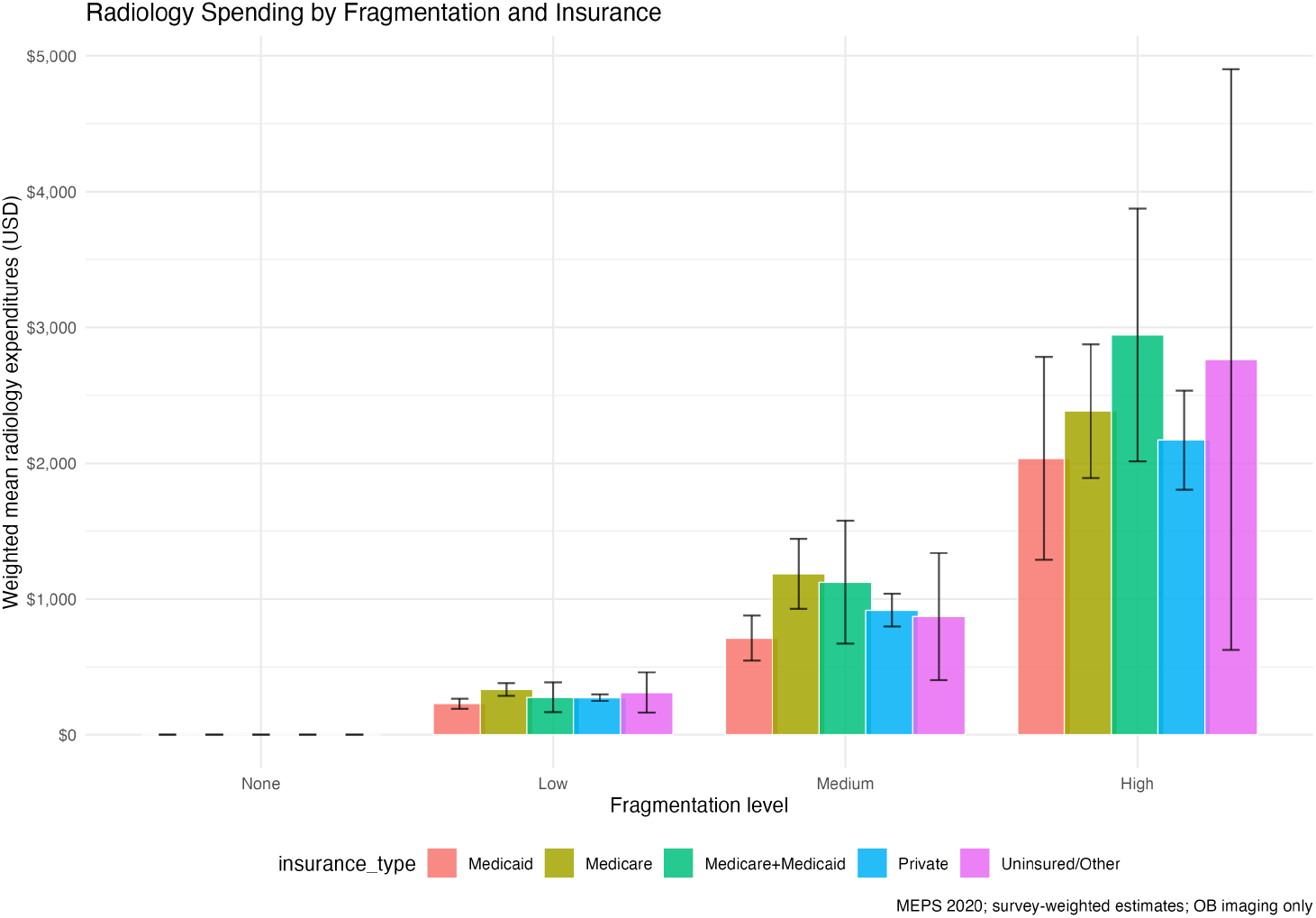
Radiology spending by fragmentation and insurance.

## 3 Results

### 3.1 Descriptive characteristics

Weighted distributions of demographic characteristics, insurance coverage, multimorbidity burden, and healthcare utilization patterns across fragmentation categories are presented in Table 1. Among adults with any office-based care, higher fragmentation levels were associated with increased overall utilization breadth and intensity across multiple care settings. The *None* fragmentation category, representing individuals with no office-based, emergency room, or inpatient encounters, had no office-based imaging by design and is included in descriptive analyses for completeness but excluded from regression models to avoid perfect prediction.

**Table 1:** Association between care fragmentation and any advanced imaging (survey-weighted logistic regression)

| Predictor | Measure | Estimate (95% CI) | *p*-value |
| --- | --- | --- | --- |
| Fragmentation: Low | Reference | — | — |
| Fragmentation: Medium vs Low | Odds Ratio | 1.42 (1.23–1.64) | < 0.001 |
| Fragmentation: High vs Low | Odds Ratio | 0.92 (0.71–1.19) | 0.54 |
| Age 50–64 vs < 50 | Odds Ratio | 1.87 (1.63–2.14) | < 0.001 |
| Age 65+ vs < 50 | Odds Ratio | 1.48 (1.11–1.99) | 0.009 |
| Female vs Male | Odds Ratio | 1.36 (1.22–1.51) | < 0.001 |
| Medicare vs Other | Odds Ratio | 1.81 (1.30–2.51) | < 0.001 |

Notes: Survey-weighted logistic regression adjusted for age group, sex, race/ethnicity, poverty status, insurance type, and total office visits. Reference category for fragmentation is Low; None excluded to avoid separation. OR = odds ratio; CI = confidence interval.

### 3.2 Association with advanced imaging

The relationship between care fragmentation and advanced imaging utilization demonstrated a non-monotonic pattern (Table 1). Compared to Low fragmentation, Medium fragmentation was associated with significantly higher odds of receiving any advanced imaging (OR 1.42, 95% CI 1.23–1.64, *p <* 0.001). However, High fragmentation did not demonstrate further increases in imaging probability (OR 0.92, 95% CI 0.71–1.19, *p* = 0.54), suggesting a plateau effect at higher fragmentation levels. Several demographic and clinical factors independently predicted advanced imaging use. Adults aged 50–64 years had substantially higher odds compared to younger adults (OR 1.87, 95% CI 1.63–2.14, *p <* 0.001), as did those aged 65 and older (OR 1.48, 95% CI 1.11–1.99, *p* = 0.009). Female sex was associated with increased imaging utilization (OR 1.36, 95% CI 1.22–1.51, *p <* 0.001), and Medicare coverage significantly predicted advanced imaging use (OR 1.81, 95% CI 1.30–2.51, *p <* 0.001).

### 3.3 Association with radiology expenditures

The two-part model revealed distinct patterns for the probability of incurring any radiology expenses versus spending levels among those with positive expenditures (Table 2, Table 3). In the first part examining any radiology spending, Medium fragmentation was associated with significantly higher odds compared to Low fragmentation (OR 1.45, 95% CI 1.25–1.69, *p <* 0.001), while High fragmentation showed no significant association (OR 0.86, 95% CI 0.65–1.14, *p* = 0.29). Medicare coverage was a strong predictor of any radiology spending (OR 1.86, 95% CI 1.33–2.59, *p <* 0.001). In the second part examining spending among those with positive expenditures, Medium fragmentation was associated with 2.36-fold higher spending (95% CI 2.04–2.72, *p <* 0.001) and High fragmentation with 3.96-fold higher spending (95% CI 3.46–4.53, *p <* 0.001) compared to Low fragmentation. Insurance type significantly modified spending patterns among those with positive expenditures.

**Table 2:**
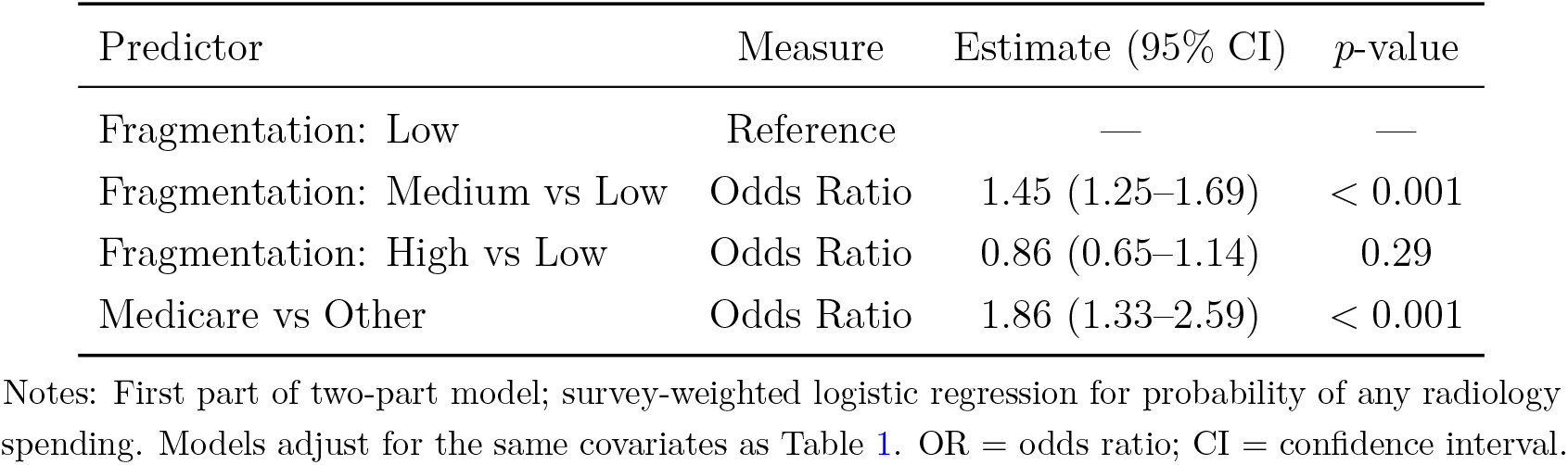
Any radiology spending (survey-weighted logistic regression)

**Table 3:** Radiology spending among spenders (survey-weighted GLM with log link, gamma family)

| Predictor | Measure |
| --- | --- |
| Fragmentation: Low | Reference |
| Fragmentation: Medium vs Low | Multiplicative Factor |
| Fragmentation: High vs Low | Multiplicative Factor |
| Medicare vs Other | Multiplicative Factor |
| Dual Medicare–Medicaid vs Other | Multiplicative Factor |
| Private vs Other | Multiplicative Factor |
| Uninsured/Other vs Private \| Multiplicative Factor \| 2.43 (1.45–4.07) \| $< 0.001$ | |

Notes: Second part of two-part model for positive radiology expenditures; generalized linear model with gamma family and log link. Estimates are exponentiated coefficients representing multiplicative effects on spending. CI = confidence interval.

**Table 4:**
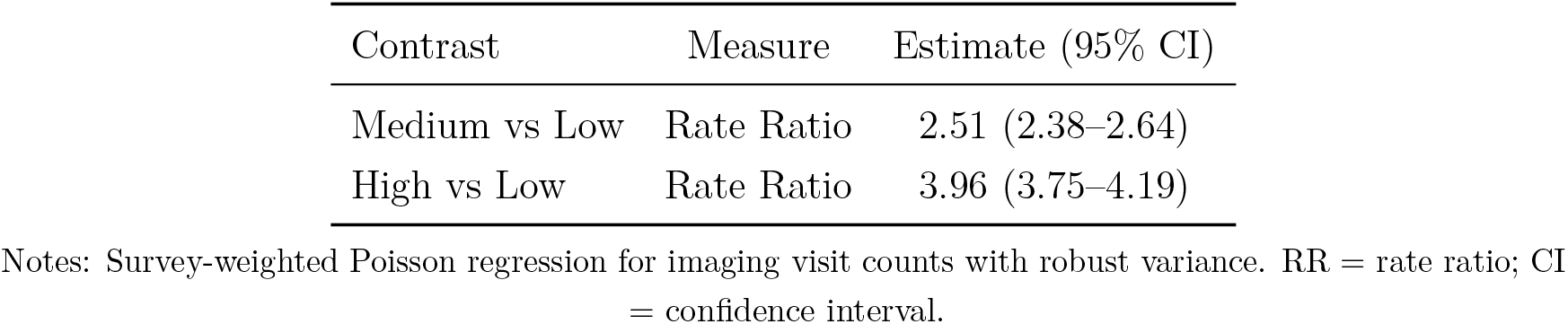
Imaging visit counts (survey-weighted Poisson regression)

| Contrast | Measure | Estimate (95% CI) |
| --- | --- | --- |
| Medium vs Low | Rate Ratio | 2.51 (2.38–2.64) |
| High vs Low | Rate Ratio | 3.96 (3.75–4.19) |

Notes: Survey-weighted Poisson regression for imaging visit counts with robust variance. RR = rate ratio; CI = confidence interval.

### 3.4 Sensitivity analyses

Sensitivity analyses confirmed the robustness of our primary findings. Alternative fragmentation cutpoints produced consistent effect directions and magnitudes. Restricting the advanced imaging outcome to MRI and CT scans only (excluding ultrasound) yielded similar qualitative patterns. Excluding the *None* fragmentation category from regression models while using Low fragmentation as the reference category avoided separation and maintained interpretability; descriptives retain *None* for completeness.

## 4 Discussion

This nationally representative study provides the first comprehensive examination of the relationship between care fragmentation and outpatient imaging utilization among U.S. adults across diverse insurance types and health status. Our findings reveal a complex relationship characterized by a threshold effect for imaging probability and a dose-response relationship for imaging intensity and expenditures.

The observed pattern where Medium fragmentation significantly increased the probability of advanced imaging (42% higher odds) while High fragmentation showed no further increase suggests a saturation effect at moderate fragmentation levels. This indicates that the transition from Low to Medium fragmentation represents a critical inflection point where imaging utilization patterns change substantially. The plateau at High fragmentation may reflect established imaging patterns among high utilizers, more conservative testing in clearly complex cases, or administrative constraints limiting further escalation.

More striking than the probability effects were the substantial increases in imaging intensity and expenditures. Among those with any radiology spending, Medium fragmentation was associated with 2.4-fold higher expenditures and High fragmentation with 4.0-fold higher expenditures compared to Low fragmentation. These effect sizes represent clinically meaningful differences with substantial economic implications. For context, the median radiology spending in our sample was $312 among those with any spending, suggesting that High fragmentation patients may incur over $1,200 in additional radiology costs annually compared to Low fragmentation patients.

Demographic and insurance factors provided additional insights into imaging utilization patterns. The strong associations with age, particularly among adults aged 50–64 years (87% higher odds) and those 65 and older (48% higher odds), likely reflect increasing prevalence of conditions requiring diagnostic imaging as well as age-related screening recommendations. The 36% higher odds among women may reflect sex-specific imaging needs, including reproductive health imaging and higher rates of certain chronic conditions requiring monitoring. Medicare coverage emerged as a particularly strong predictor of both imaging probability (81% higher odds) and expenditures, consistent with previous research.

### 4.1 Implications

Clinically, these results emphasize the need for enhanced continuity of care and targeted care coordination interventions for patients experiencing Medium and High levels of care fragmentation. Clinicians should be vigilant about potentially unnecessary imaging in patients who receive care across multiple providers and settings, implementing systematic approaches to track imaging history and avoid duplication. From a policy standpoint, these findings support reimbursement models and programmatic interventions designed to reduce care fragmentation, such as patient-centered medical homes, accountable care organizations, and care coordination payment incentives. Health systems and payers should consider monitoring imaging volume and expenditures among high-fragmentation patient subgroups as quality and cost-containment measures.

### 4.2 Strengths and limitations

The use of nationally representative MEPS data with appropriate survey weighting ensures generalizability to the U.S. adult population, representing over 251 million adults. The fragmentation measure, while simplified for administrative data, provides a transparent and feasible proxy that can be implemented using standard utilization data. The analytical approach combining both probability and intensity outcomes provides a comprehensive view of the fragmentation–imaging relationship. However, limitations include the lack of provider-level identifiers, focus on office-based imaging (potentially underestimating total imaging), cross-sectional design precluding causal inference, potential residual confounding, reliance on patient-reported service indicators, and the unique context of 2020 during the COVID-19 pandemic.

## 5 Conclusions

Care fragmentation is significantly associated with increased advanced imaging utilization and substantially higher radiology expenditures among U.S. adults. The relationship demonstrates a complex pattern where Medium fragmentation increases both the probability of receiving advanced imaging and spending levels, while High fragmentation shows plateauing probability but continued increases in spending intensity. Interventions targeting care coordination and continuity may reduce potentially unnecessary imaging utilization and associated costs while maintaining appropriate access to needed diagnostic services. Future research should examine the clinical appropriateness of imaging in fragmented versus coordinated care settings and evaluate the effectiveness of specific interventions designed to improve care coordination and reduce fragmentation-related overuse.

## Data Availability

All data produced in the present study are available upon reasonable request to the authors.

## 6 Data availability

This study uses publicly available MEPS 2020 public-use files from AHRQ (HC-224, HC-220G, HC-220IF1, HC-222). Analysis code and derived, non-identifiable outputs are available upon reasonable request.

## 7 Ethics approval and consent

MEPS public-use data are de-identified; this secondary analysis does not require IRB review. No individual consent was required.

## 8 Funding

No specific funding was received for this work.

## 9 Author contributions

Andrew Bouras: conception and design; data curation; analysis; drafting. Dhruv Patel: interpretation; critical revision. All authors approved the final manuscript.

## 10 Acknowledgments

We thank the Agency for Healthcare Research and Quality (AHRQ) for MEPS data access and the MEPS team for documentation and support.

## Abbreviations

MEPS: Medical Expenditure Panel Survey
FYC: Full-Year Consolidated
FOB: Office-Based
FER: Emergency Room
FMRI: Magnetic Resonance Imaging
FCT: Computed Tomography
FPSU: Primary Sampling Unit
FOR: Odds Ratio
FRR: Rate Ratio

## 12 Supplementary material

Supplementary tables and extended methods (e.g., alternative fragmentation cutpoints; MRI/CT-only outcome) are available upon request.

## Notes

### Competing Interest Statement

The authors have declared no competing interest.

### Funding Statement

This study did not receive any funding

### Author Declarations

The study used ONLY openly available human data that were publicly accessible before the initiation of the study. Data were obtained from the Medical Expenditure Panel Survey (MEPS) public-use files hosted by the Agency for Healthcare Research and Quality (AHRQ). No registration, application, screening, or data use agreement was required. All analyses were conducted on the public-use files and documentation available at the following download pages: MEPS HC-224: 2020 Full-Year Consolidated Data File - https://meps.ahrq.gov/data_stats/download_data/pufs/h224/h224doc.shtml MEPS HC-220G: 2020 Office-Based Medical Provider Visits - https://meps.ahrq.gov/data_stats/download_data/pufs/h220g/h220gdoc.shtml MEPS HC-220IF1: 2020 Condition-Event Link - https://meps.ahrq.gov/data_stats/download_data/pufs/h220if1/h220if1doc.shtml MEPS HC-222: 2020 Medical Conditions - https://meps.ahrq.gov/data_stats/download_data/pufs/h222/h222doc.shtml Data can be located by visiting the above pages and downloading the corresponding public-use files and codebooks.

## References

[1] Kern LM, Seirup JK, Rajan M, et al. Fragmented ambulatory care and subsequent healthcare utilization among Medicare beneficiaries. Am J Manag Care. 2018;24(9):e278–e284.

[2] Romano MJ, Segal JB, Pollack CE. The association between continuity of care and the overuse of medical procedures. JAMA Intern Med. 2015;175(7):1148–1154.

[3] Prior A, Vestergaard CH, Davydow DS, et al. Healthcare fragmentation and the risk of mortality: a longitudinal study. BMC Med. 2023;21(1):201.

[4] Pollack CE, Hussey PS, Rudin RS, et al. Measuring care continuity: a comparison of claims-based methods. Med Care. 2016;54(5):e30–e34.

[5] Iglehart JK. Health insurers and medical-imaging policy—a work in progress. N Engl J Med. 2009;360(10):1030–1037.

[6] Smith-Bindman R, Miglioretti DL, Johnson E, et al. Use of diagnostic imaging studies and associated radiation exposure for patients enrolled in large integrated health care systems, 1996–2010. JAMA. 2012;307(22):2400–2409.

[7] Lang K, Huang H, Lee DW, et al. National trends in advanced outpatient diagnostic imaging utilization: an analysis of the medical expenditure panel survey, 2000–2009. BMC Med Imaging. 2013;13:40.

[8] Levin DC, Rao VM, Parker L, et al. Recent trends in imaging utilization in hospital settings: implications for future planning. J Am Coll Radiol. 2019;16(4 Pt A):331–337.

[9] Medicare Payment Advisory Commission. Report to the Congress: Medicare and the health care delivery system. Washington, DC: MedPAC; 2021.

[10] McDonald KM, Sundaram V, Bravata DM, et al. Closing the quality gap: a critical analysis of quality improvement strategies (Vol. 7: Care coordination). Rockville, MD: Agency for Healthcare Research and Quality; 2007.

[11] Bodenheimer T. Coordinating care—a perilous journey through the health care system. N Engl J Med. 2008;358(10):1064–1071.

[12] Studdert DM, Mello MM, Sage WM, et al. Defensive medicine among high-risk specialist physicians in a volatile malpractice environment. JAMA. 2005;293(21):2609–2617.

[13] Bach PB, Pham HH, Schrag D, et al. Primary care physicians who treat blacks and whites. N Engl J Med. 2004;351(6):575–584.

[14] Frandsen BR, Joynt KE, Rebitzer JB, et al. Care fragmentation, quality, and costs among chronically ill patients. Am J Manag Care. 2015;21(5):355–362.

[15] Nyweide DJ, Bynum JP. Relationship between continuity of ambulatory care and risk of emergency department episodes among older adults. Ann Emerg Med. 2017;69(4):407– 415.

[16] MEPS HC-224 2020 Full Year Consolidated Data File. Agency for Healthcare Research and Quality. https://meps.ahrq.gov/data_stats/download_data/pufs/h224/h224doc.shtml

[17] MEPS Show Cards (imaging categories). Medical Expenditure Panel Survey. https://www.mepsdocs.org/Home/Showcards

